# Clinical Characteristics of Two Human to Human Transmitted Coronaviruses: Corona Virus Disease 2019 versus Middle East Respiratory Syndrome Coronavirus

**DOI:** 10.1101/2020.03.08.20032821

**Authors:** Ping Xu, Guo-Dong Sun, Zhi-Zhong Li

## Abstract

After the outbreak of the middle east respiratory syndrome (MERS) worldwide in 2012. Currently, a novel human coronavirus has caused a major disease outbreak, and named corona virus disease 2019 (COVID-19). The emergency of MRES-COV and COVID-19 has caused global panic and threatened health security. Unfortunately, the similarities and differences between the two coronavirus diseases remain to be unknown. The aim of this study, therefore, is to perform a systematic review to compare epidemiological, clinical and laboratory features of COVID-19 and MERS-COV population. We searched PubMed, EMBASE and Cochrane Register of Controlled Trials database to identify potential studies reported COVID-19 or MERS-COV. Epidemiological, clinical and laboratory outcomes, the admission rate of intensive cure unit (ICU), discharge rate and fatality rate were evaluated using GraphPad Prism software. Thirty-two studies involving 3770 patients (COVID-19 = 1062, MERS-COV = 2708) were included in this study. The present study revealed that compared with COVID-19 population, MERS-COV population had a higher rate of ICU admission, discharge and fatality and longer incubation time. It pointed out that fever, cough and generalised weakness and myalgia were main clinical manifestations of both COVID-19 and MERS-COV, whereas ARDS was main complication. The most effective drug for MERS-COV is ribavirin and interferon.

## Introduction

Coronaviruses are RNA viruses with envelope and non-segmented positive-sense, causing respiratory and intestinal tract infections in humans and other mammals [1]. Despite majority of patients present mild symptoms and good prognosis, the spread of two β-coronaviruses, severe acute respiratory syndrome coronavirus (SARS-COV) and middle east respiratory syndrome coronavirus (MERS-COV), have resulted in the infection of thousands of individuals in past decades, with the fatality rate of 10% for SARS-COV population and 37% for MERS-COV population [2-4]. Before 2019, there is a total of six identified coronaviruses, but that might only be a small part of the iceberg, with potentially more novel and terrible zoonoses to be presented.

As December 8, 2019, a series of unexplained pneumonia cases linked to Huanan seafood wholesale market has been reported in Wuhan, Hubei, China [5-6]. Clinical characteristics of this pneumonia were very similar to those of viral pneumonia such as MERS-COV, initially severe acute respiratory infection, followed by developing rapidly acute respiratory distress syndrome (ARDS) and even acute respiratory failure [7]. Sequencing data from throat swab samples of a patient indicated a novel coronavirus subsequently named corona virus disease 2019 (COVID-19) by world health organization (WHO). So far, more than 70000 confirmed cumulative cases have been reported in China, and more than 10000 confirmed cumulative cases in other countries such as South Korea, Japan, Italy, Iran and the USA.

Although many previous studies have reported clinical characteristics of COVID-19 or MERS-COV diseases [8-11], systematic comparison of clinical features between COVID-19 and MERS-COV diseases has yet been published. Thus, the purpose of this study is to perform a systematic review of epidemiological, clinical and laboratory characteristics of patients infected by COVID-19 or MERS-COV disease, and to compare COVID-19 and MERS-COV in the context of their incubation, laboratory features, admission rate of intensive cure unit (ICU) and rate of discharge and fatality, which will provide a comprehensive reference for clinical physicians in treatment of coronavirus diseases.

## Material and Methods

### Search strategy

A comprehensive and systematic search was performed using PubMed, EMBASE and Cochrane Register of Controlled Trials database up to 26 February 2020. Medical Subject Heading (MSH) terms and keywords were used so as to retrieve as many potential documents as possible. Terms for MERS-COV included: Middle East Respiratory Syndrome Coronavirus [Mesh] OR MERS-CoV OR MERS Virus OR MERS Viruses OR Virus, MERS OR Viruses, MERS OR Middle East respiratory syndrome-related coronavirus OR Middle East respiratory syndrome related coronavirus. They were combined with terms specifying COVID-19: ((Wuhan coronavirus) OR (Wuhan seafood market pneumonia virus) OR (SARS2) OR (COVID-19 virus) OR (coronavirus disease 2019 virus) OR (SARS-CoV-2) OR (2019-nCoV) OR (2019 novel coronavirus)). If necessary, we also contacted corresponding author to obtain accurate data.

### Eligibility criteria

The study that met following criteria were included: (1) reporting clinical characteristics of COVID-19 or MERS-COV disease, (2) minimum sample size of five, (3) confirmed COVID-19 or MERS-COV disease, (4) English literature. Studies were excluded as following criteria: duplicate publications, case report, meta-analysis, letter, review, technology report, commentary, animal trial, correspondence, predictive study, guidence, radiograph study and meeting report.

### Study Selection

At the beginning, 4743 potential publications were identified. We removed 678 duplicates and reviewed the titles and abstracts of remaining 4065 publications. 4015 publications were excluded for following reasons: not involved research point (n = 1378), review (n = 569), no English (n = 33), case report (n = 316), meta-analysis (n = 1), letter (n = 127), technology report (n = 196), commentaries (n = 61), animal study (n = 1107), correspondence (n = 29), predictive study (n = 110), guidence (n = 28), meeting report (n = 16) and radiograph (n = 44). Then, a comprehensive review of full-text was conducted for remaining 50 publications. Two reviewers independently screened eligible literature, and any argument was solved by discussion with a third reviewer. Finally, thirty-two studies were included in this study [8-39]. The process was shown in Figure 1.

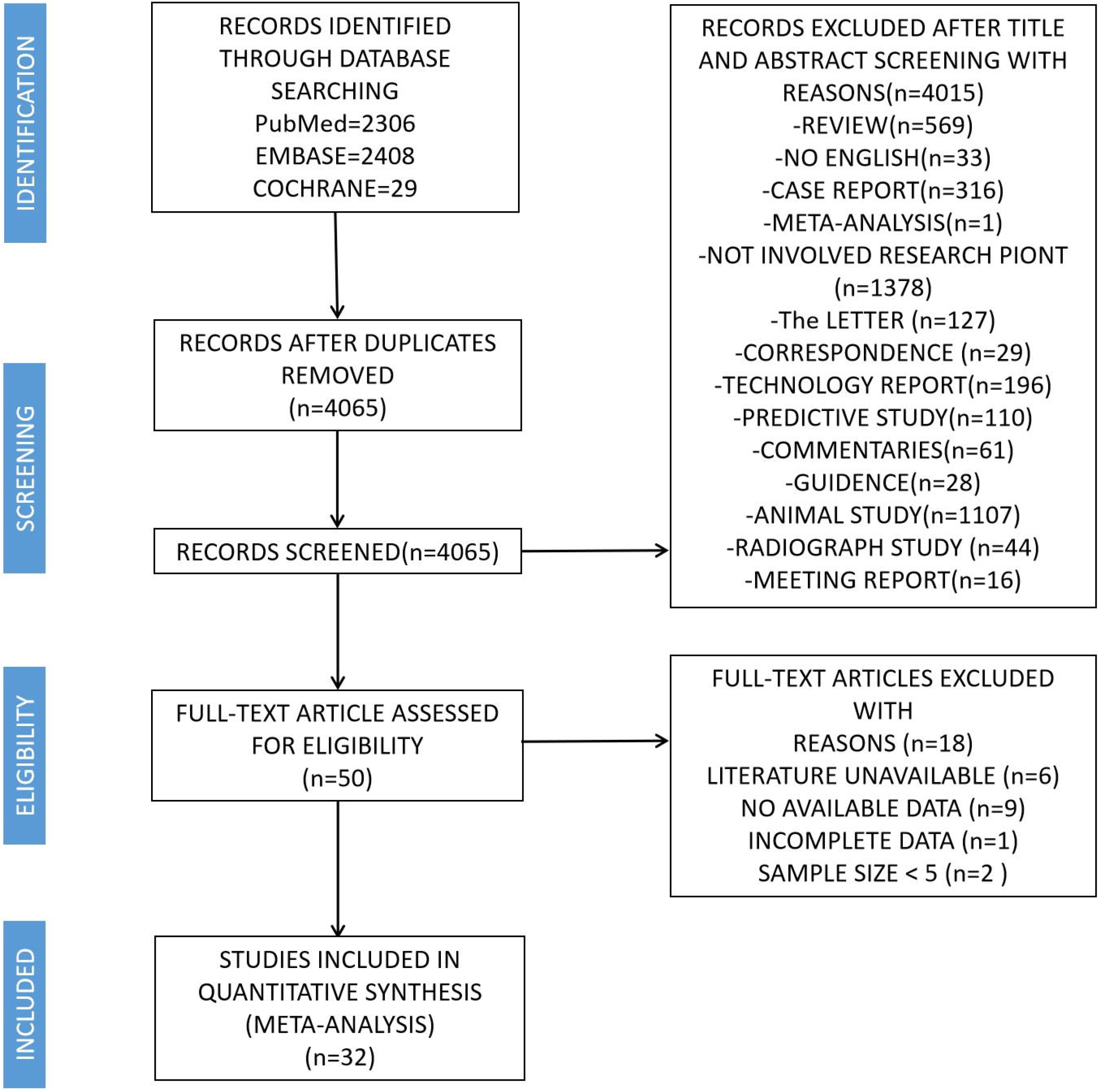

### Data Extraction

Two reviewers extracted independently common, clinical and laboratory characteristics of included studies, with disagreements were solved by discussion with a third reviewer. The extracted data included incubation time, white blood cell (WBC) count, lymphocyte count, c-reactive protein (CRP), alamine aminotransferase (ALT), aspartate aminotransferase (AST), creatinine, creatine kinase (CK), the admission rate of intensive cure unit (ICU), rate of discharge and fatality, symptom, comorbidity, complications and cure rate of drug. For normality distribution data, outcomes were extracted directly. For skewness distribution data, the outcomes was extracted after being converted as specific formula [40].

### Quality Assessment

The quality assessment of included studies was performed through the Newcastle-Ottawa Quality Assessment Scale (NOS), as recommended by the Cochrane Non-Randomized Studies [41]. The NOS includes three parts for risk of bias, with nine points in total: (1) selection of research groups (four points); (2) inter-group comparability (two points); and (3) ascertainment of exposure and outcomes (three points) for case–control and cohort studies, respectively. Study that scored 6 or more was qualified for systematic review [42]. The assessment process was completed by two reviewers independently. All debates were solved by discussion with a third reviewer.

### Statistical Analysis

All statistical analyses and graphs were generated and plotted using GraphPad Prism version 7.00 software (GraphPad Software Inc). The p value < 0.05 suggests significant difference.

## Results

### Study Characteristics

All included studies were retrospective study [8-39]. Among ten studies reported COVID-19, one trial was performed in Netherlands [12] and remaining nine trials were conducted in China [8-9, 13-16, 37-39]. The sample size ranged from 6 to 425, and had a total of 1062 patients (male = 610, female = 452). The year of publications was in 2020. On the other hand, among twenty-two studies reported MERS-COV, fifteen studies were conducted in Saudi Arabia [11, 17-18, 20-23, 27-30, 32, 34-36], five trials were performed in South Korea [24-26, 31, 33], one trial was performed in Iran [19] and one trial was conducted in Japan [10]. The sample size ranged from 5 to 883, and had a total of 2708 patients. The year of publications ranged from 2013 to 2020. Clinical and laboratory characteristics of included study were shown in Table 1 and Table 2 respectively.

**Table.1.**
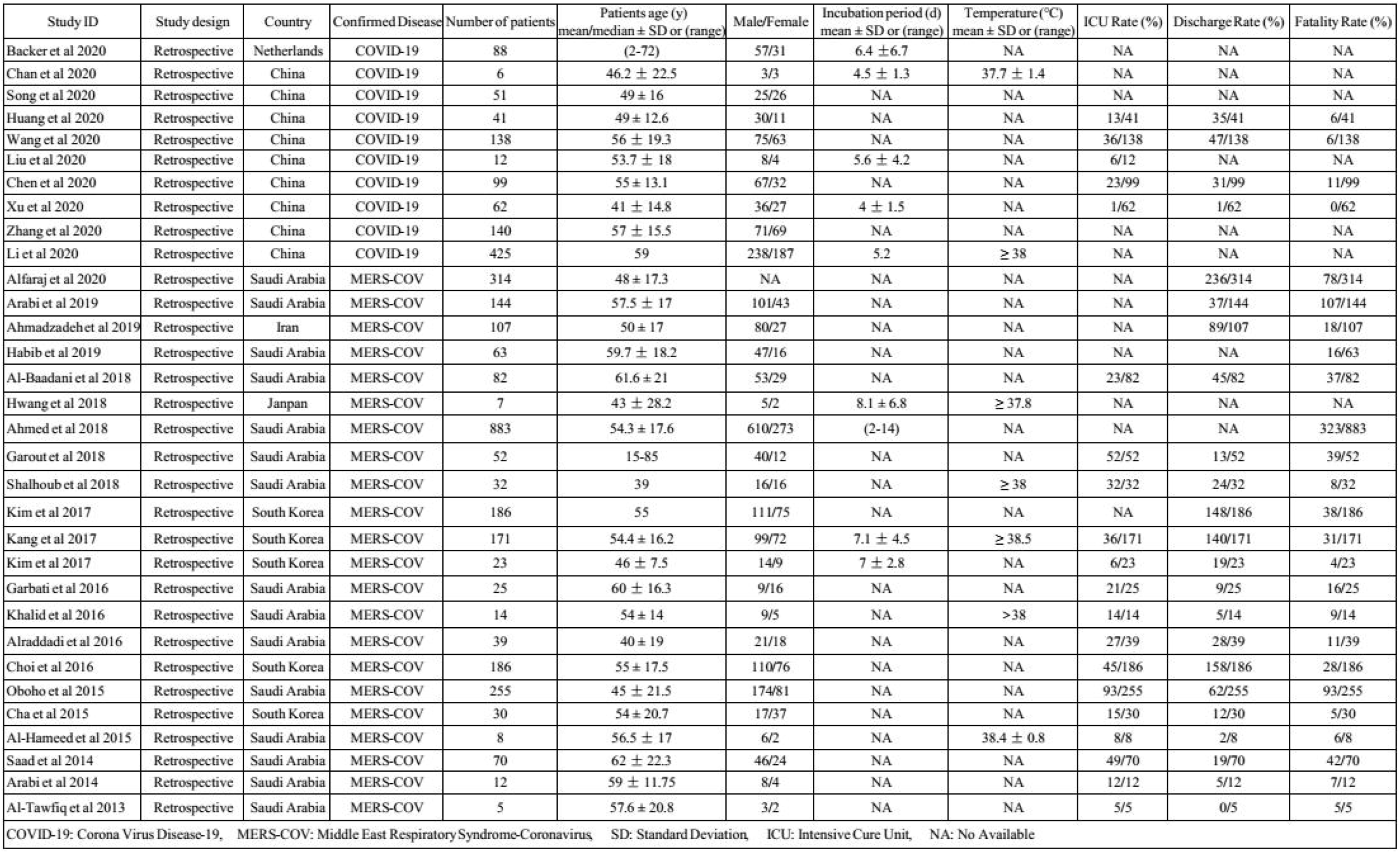
Common and Clinical Characteristics of Included Studies.

**Table.2.**
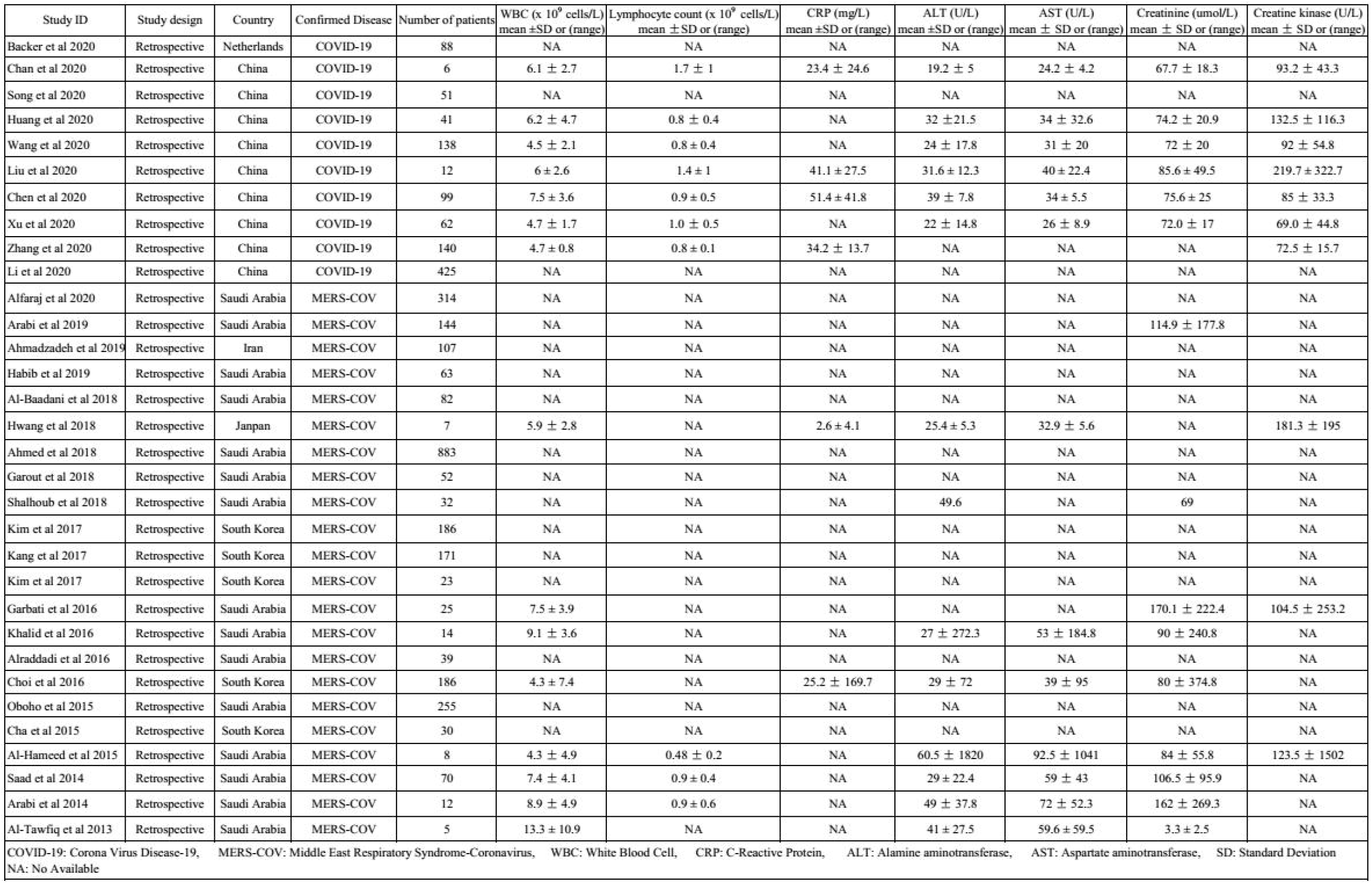
Laboratary Outcomes of Included Studies.

### Quality Assessment

Among thirty-two included studies, four studies obtained 6 points of NOS [26, 28, 32, 35], and remaining twenty-eight studies obtained 7 points of NOS or more [8-25, 27, 29-31, 33-34, 36-39]. The result of quality assessment was presented in Table 3.

**Table3.** Quality Assessment of Included Studies.

| Study | Selection | Comparability | Outcome | Total scores |
| --- | --- | --- | --- | --- |
| Backer et al 2020 | 4 | 1 | 2 | 7 |
| Chan et al 2020 | 4 | 2 | 2 | 8 |
| Song et al 2020 | 4 | 2 | 2 | 8 |
| Huang et al 2020 | 4 | 2 | 2 | 8 |
| Wang et al 2020 | 4 | 2 | 2 | 8 |
| Liu et al 2020 | 4 | 1 | 2 | 7 |
| Li et al 2020 | 3 | 2 | 2 | 7 |
| Chen et al 2020 | 4 | 1 | 2 | 7 |
| Xu et al 2020 | 4 | 1 | 2 | 7 |
| Zhang et al 2020 | 4 | 1 | 2 | 7 |
| Alfaraj et al 2020 | 4 | 2 | 2 | 8 |
| Arabi et al 2019 | 4 | 2 | 2 | 8 |
| Ahmadzadeh et al 2019 | 4 | 2 | 2 | 8 |
| Habib et al 2019 | 4 | 1 | 2 | 7 |
| Al-Baadani et al 2018 | 4 | 2 | 2 | 8 |
| Hwang et al 2018 | 4 | 2 | 2 | 8 |
| Ahmed et al 2018 | 4 | 2 | 2 | 8 |
| Garout et al 2018 | 4 | 2 | 2 | 8 |
| Shalhoub et al 2018 | 4 | 1 | 2 | 7 |
| Kim et al 2017 | 4 | 1 | 2 | 7 |
| Kang et al 2017 | 4 | 2 | 2 | 8 |
| Kim et al 2017 | 3 | 1 | 2 | 6 |
| Garbati et al 2016 | 4 | 1 | 2 | 7 |
| Khalid et al 2016 | 4 | 1 | 2 | 7 |
| Alraddadi et al 2016 | 4 | 1 | 2 | 7 |
| Choi et al 2016 | 4 | 1 | 2 | 7 |
| Al-Hameed et al 2015 | 3 | 1 | 2 | 6 |
| Oboho et al 2015 | 3 | 1 | 2 | 6 |
| Cha et al 2015 | 4 | 1 | 2 | 7 |
| Saad et al 2014 | 4 | 1 | 2 | 7 |
| Arabi et al 2014 | 4 | 1 | 2 | 7 |
| Al-Tawfiq et al 2013 | 3 | 1 | 2 | 6 |

### Clinical Symptom

For COVID-19 population, the number of patients with fever was 480 (45.2%), cough was 373 (35.1%), generalised weakness and myalgia was 318 (29.9%), stuffy or rhinorrhea was 6 (0.6%), pharyngalgia was 32 (3%), chest pain was 10 (0.9%), diarrhoea or anorexia was 123 (11.6%), dyspnoea was 140 (13.2%) and dizziness or headache was 60 (5.6%). For MERS-COV population, the amount of patients with fever was 404 (14.9%), cough was 424 (15.7%), generalised weakness and myalgia was 337 (12.4%), stuffy or rhinorrhoea was 17 (0.6%), pharyngalgia was 47 (1.7%), chest pain was 27 (1%), diarrhoea or anorexia was 128 (4.7%), dyspnoea was 271 (10%) and dizziness or headache was 131 (4.8%). The Above results were shown in Table 4.

**Table 4.** Description of Clinical Symptoms in COVID-19 and MERS-COV Populations.

| Symptom | COVID-19, No (n/total %) | MERS-COV, No (n/total %) |
| --- | --- | --- |
| Fever | 480 (45.2%) | 404 (14.9%) |
| Cough | 373 (35.1%) | 424 (15.7%) |
| Generalised weakness and myalgia | 318 (29.9%) | 337 (12.4%) |
| Stuffy and Rhinorrhoea | 6 (0.6%) | 17 (0.6%) |
| Pharyngalgia | 32 (3%) | 47 (1.7%) |
| Chest pain | 10 (0.9%) | 27 (1%) |
| Diarrhoea or Anorexia | 123 (11.6%) | 128 (4.7%) |
| Dyspnoea | 140 (13.2%) | 271 (10%) |
| Dizziness or headache | 60 (5.6%) | 131 (4.8%) |

COVID-19: Corona Virus Disease-19, MERS-COV: Middle East Respiratory Syndrome-Coronavirus Disease

### Complications

For COVID-19 population, the main complications included shock, arrhythmia, acute respiratory distress syndrome (ARDS), acute cardiac injury, acute kidney injury and acute liver injury, and the amount of which was 17 (1.6%), 28 (2.6%), 51 (4.8%), 11 (1%), 10 (0.9%) and 2 (0.2%) respectively. For MERS-COV population, the number of individuals presented shock was 22 (0.8%), arrhythmia was 11 (0.4%), ARDS was 83 (3.1%), acute cardiac injury 10 (0.4%), acute kidney injury was 30 (1.1%), acute liver injury was 22 (0.8%) and neurological symptoms was 4 (0.1%). The results were shown in Table 5.

**Table 5.** Distribution of Complications in COVID-19 and MERS-COV Populations.

| Complication | COVID-19, No (n/total %) | MERS-COV, No (n/total %) |
| --- | --- | --- |
| Shock | 17 (1.6%) | 22 (0.8%) |
| Arrhythmia | 28 (2.6%) | 11 (0.4%) |
| ARDS | 51 (4.8%) | 83 (3.1%) |
| Acute cardiac injury | 11 (1%) | 10 (0.4%) |
| Acute kidney injury | 10 (0.9%) | 30 (1.1%) |
| Acute liver injury | 2 (0.2%) | 22 (0.8%) |
| Neurological symptoms | 0 | 4 (0.1%) |

COVID-19: Corona Virus Disease-19, MERS-COV: Middle East Respiratory Syndrome-Coronavirus Disease ARDS : Acute Respiratory Distress Syndrome

### Comorbidity

Among COVID-19 population, 43 (4%) patients had diabetes, 101 (9.5%) patients had hypertension, 77 (7.3%) patients had cardiovascular disease, 10 (0.9%) patients had chronic obstructive pulmonary disease (COPD), 19 (1.8%) patients had malignancy, 24 (2.3%) patients had chronic liver disease, 13 (1.2%) patients had cerebrovascular disease and 5 (0.5%) patients had chronic kidney disease. Among MERS-COV population, the number of patients with diabetes was 247 (9.1%), hypertension was 244 (9%), cardiovascular disease was 161 (5.9%), COPD was 83 (3.1%), malignancy was 70 (2.6%), chronic liver disease was 28 (1%), cerebrovascular disease was 47 (1.7%), chronic kidney disease was 167 (6.2%) and obesity was 42 (1.6%). Those results were shown in Table 6.

**Table 6.** Distribution of Comorbidities in COVID-19 and MERS-COV Populations.

| Comorbidity | COVID-19, No (n/total %) | MERS-COV, No (n/total %) |
| --- | --- | --- |
| Diabetes | 43 (4%) | 247 (9.1%) |
| Hypertension | 101 (9.5%) | 244 ( 9%) |
| Cardiovascular disease | 77 (7.3%) | 161 (5.9%) |
| Chronic obstructive pulmonary disease | 10 (0.9%) | 83 (3.1%) |
| Malignancy | 19 (1.8%) | 70 (2.6%) |
| Chronic liver disease | 24 (2.3%) | 28 (1%) |
| Cerebrovascular disease | 13 (1.2%) | 47 (1.7%) |
| Chronic kidney disease | 5 ( 0.5%) | 167 (6.2%) |
| Obesity | 0 | 42 (1.6%) |
| COVID-19: Corona Virus Disease-19, MERS-COV: Middle East Respiratory Syndrome-Coronavirus |  |  |

### Clinical and Laboratory Characteristics of COVID-19 versus MERS-COV

Systematic review was performed for clinical and laboratory outcomes of coronavirus disease. There was no significant difference in age (50.9 ± 2 vs. 53.6 ± 1.5, P = 0.3), WBC (5.7 ± 0.4 vs. 7.6 ± 1, P = 0.1), lymphocyte count (1.1 ± 0.1 vs. 0.8 ± 0.1, P = 0.2), CRP (37.6 ± 5.8 vs. 13.9 ± 11.3, P = 0.1), ALT (28 ± 3.1 vs. 37.3 ± 5.1, P = 0.2), creatinine (74.5 ± 0.4 vs. 101.4 ± 18.5, P = 0.2) and creatine kinase (109.1 ± 20 vs. 136.4 ± 23.1, P = 0.5) between COVID-19 and MERS-COV population. Compared with COVID-19 population, a higher AST (31.5 ± 2.4 vs. 58.3 ± 7.6, P = 0.009) and longer incubtion time (5.1 ± 0.5 vs. 7.4 ± 0.4, P = 0.02) were found in MERS-COV population. The results were shown in Table 7.

**Table 7.** Clinical and Laboratory Characteristics of COVID-19 versus MERS-COV.

| Characteristics | COVID-19<br>Mean ± SEM | MERS-COV<br>Mean ± SEM | P value |
| --- | --- | --- | --- |
| Age (y) | 50.9 ± 2 | 53.6 ± 1.5 | 0.3 |
| Incubtion time (d)* | 5.1 ± 0.5 | 7.4 ± 0.4 | 0.02 |
| WBC (x 10<sup>9</sup> cells/L) | 5.7 ± 0.4 | 7.6 ± 1 | 0.1 |
| Lymphocyte count (x 10<sup>9</sup> cells/L) | 1.1 ± 0.1 | 0.8 ± 0.1 | 0.2 |
| CRP (mg/L) | 37.6 ± 5.8 | 13.9 ± 11.3 | 0.1 |
| ALT (U/L) | 28 ± 3.1 | 37.3 ± 5.1 | 0.2 |
| AST (U/L)* | 31.5 ± 2.4 | 58.3 ± 7.6 | 0.009 |
| Creatinine (umol/L) | 74.5 ± 2.5 | 101.4 ± 18.5 | 0.2 |
| Creatine kinase (U/L) | 109.1 ± 20 | 136.4 ± 23.1 | 0.5 |

COVID-19: Corona Virus Disease-19, MERS-COV: Middle East Respiratory Syndrome-Coronavirus, WBC: White Blood Cell, CRP: C-Reactive Protein, ALT: Alamine aminotransferase, AST: Aspartate aminotransferase, SEM: Standard Eerror of Mean, \*: Significant Difference

### Incidence of ICU Admission, Discharge and Fatality

The higher admission rate of ICU was found in MERS-COV population than in COVID-19 population (43.6% vs. 22.4%, Figure 2). There was a higher discharge rate in MERS-COV populations compared with COVID-19 population (59.9% vs. 33.5%, Figure 3). The lower fatality rate was found in COVID-19 population compared with MERS-COV population (6.8% vs. 34.1%, Figure 4).

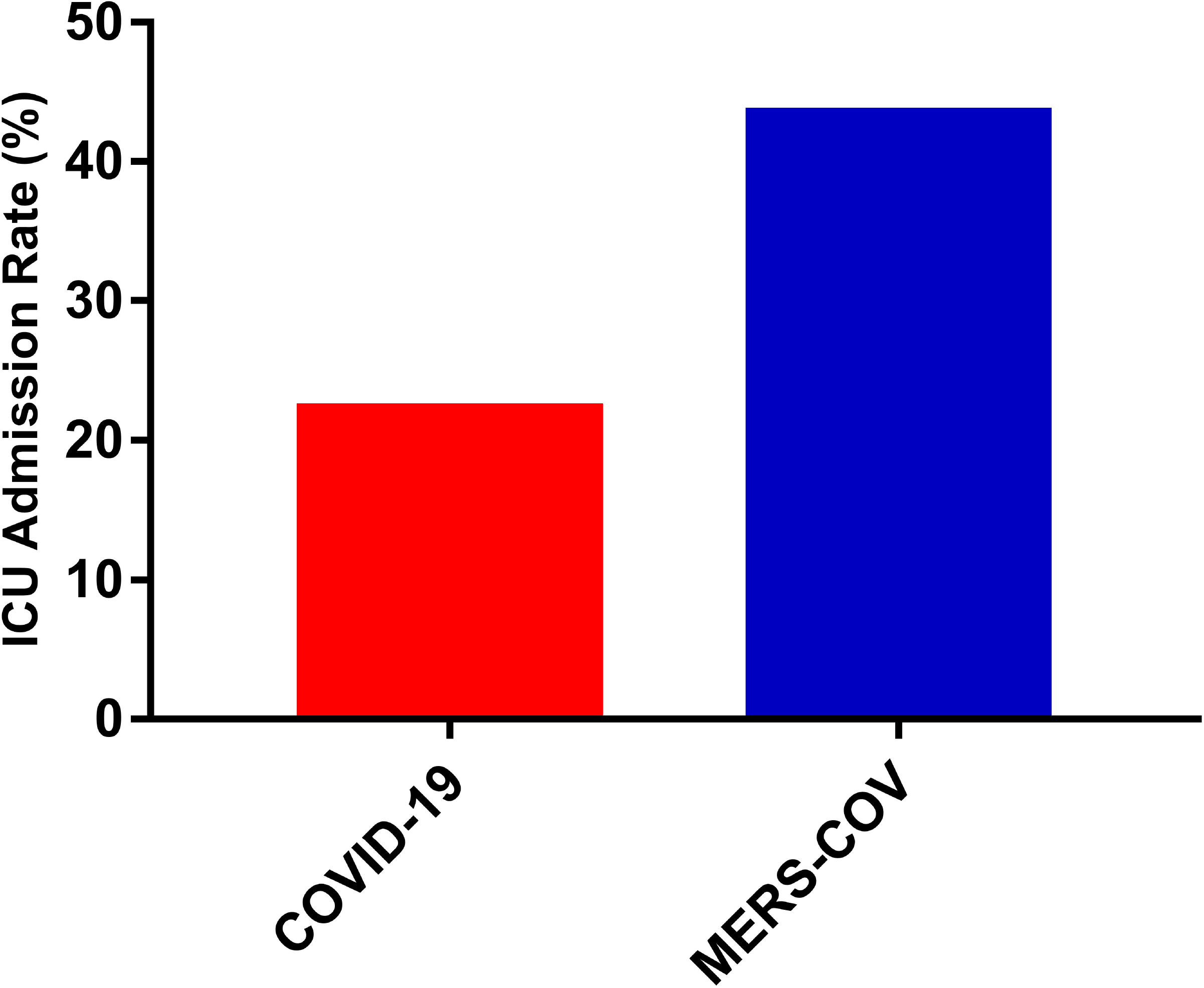

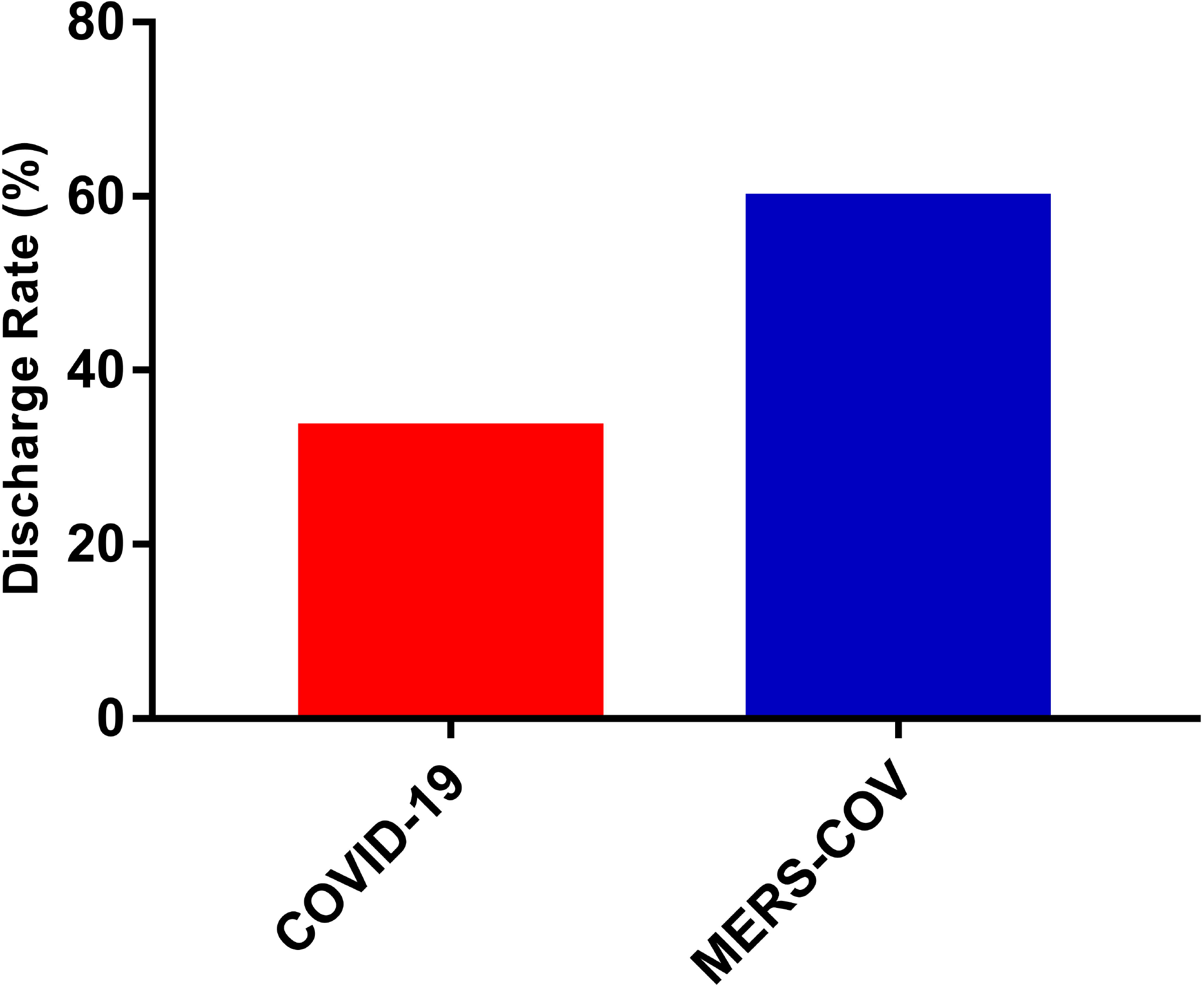

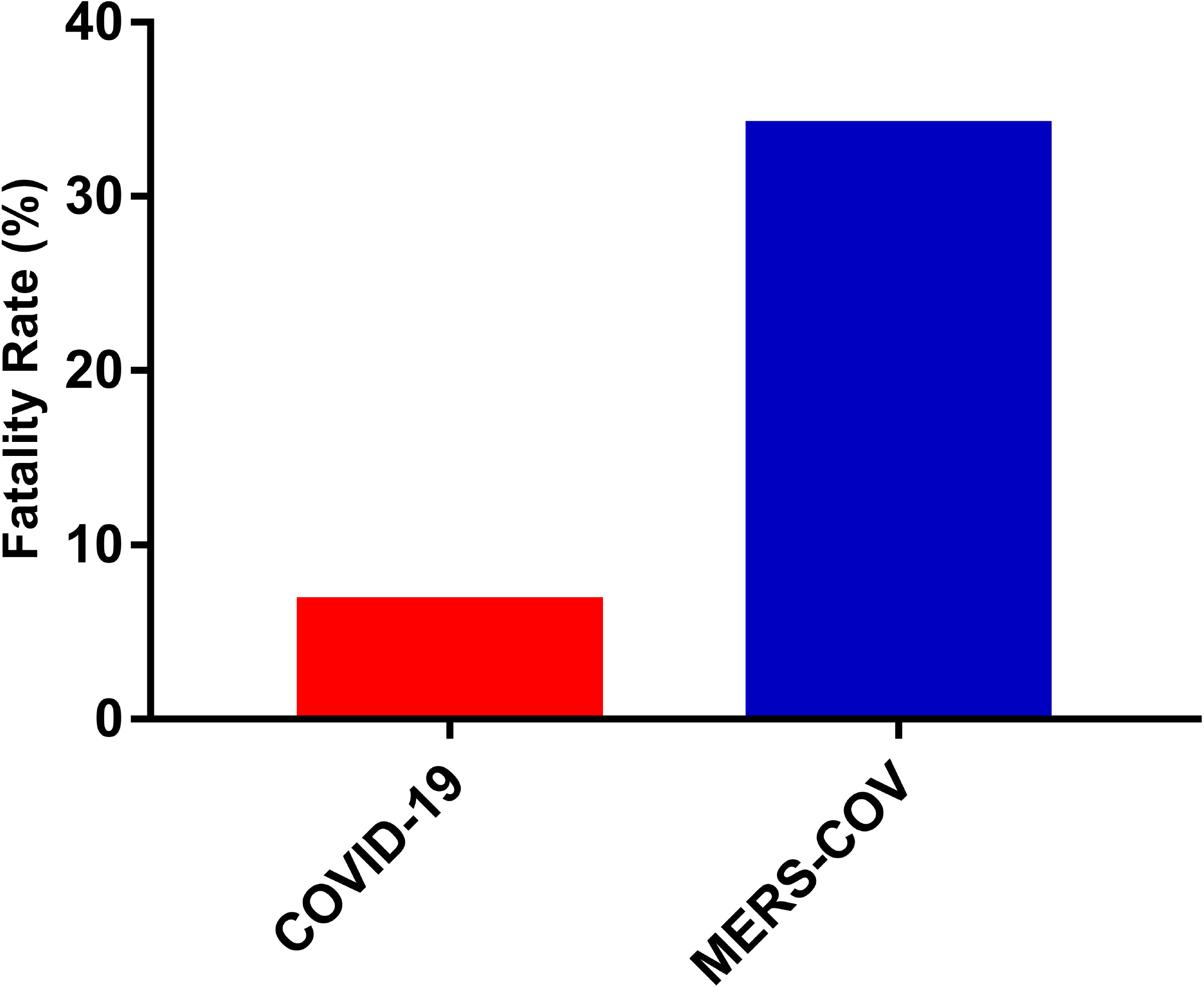

### Cure rate of drugs for MERS-COV

Of 93 patients treated by ribavirin and interferon, 67 (74.2%) were cured. Of 39 patients treated by oseltamivir, 27 (69.2%) were cured. Of 70 patients treated by antivirals, 47 (67.1%) were cured. Of 8 patients treated by intravenous immunoglobulin, 5 (62.5%) were cured. Of 9 patients treated by ribavirin and lopinavir/ritonavir, 4 (44.4%) were cured. Of 18 patients treated by corticosteroids, 7 (38.9%) were cured. The results were shown in Figure 5.

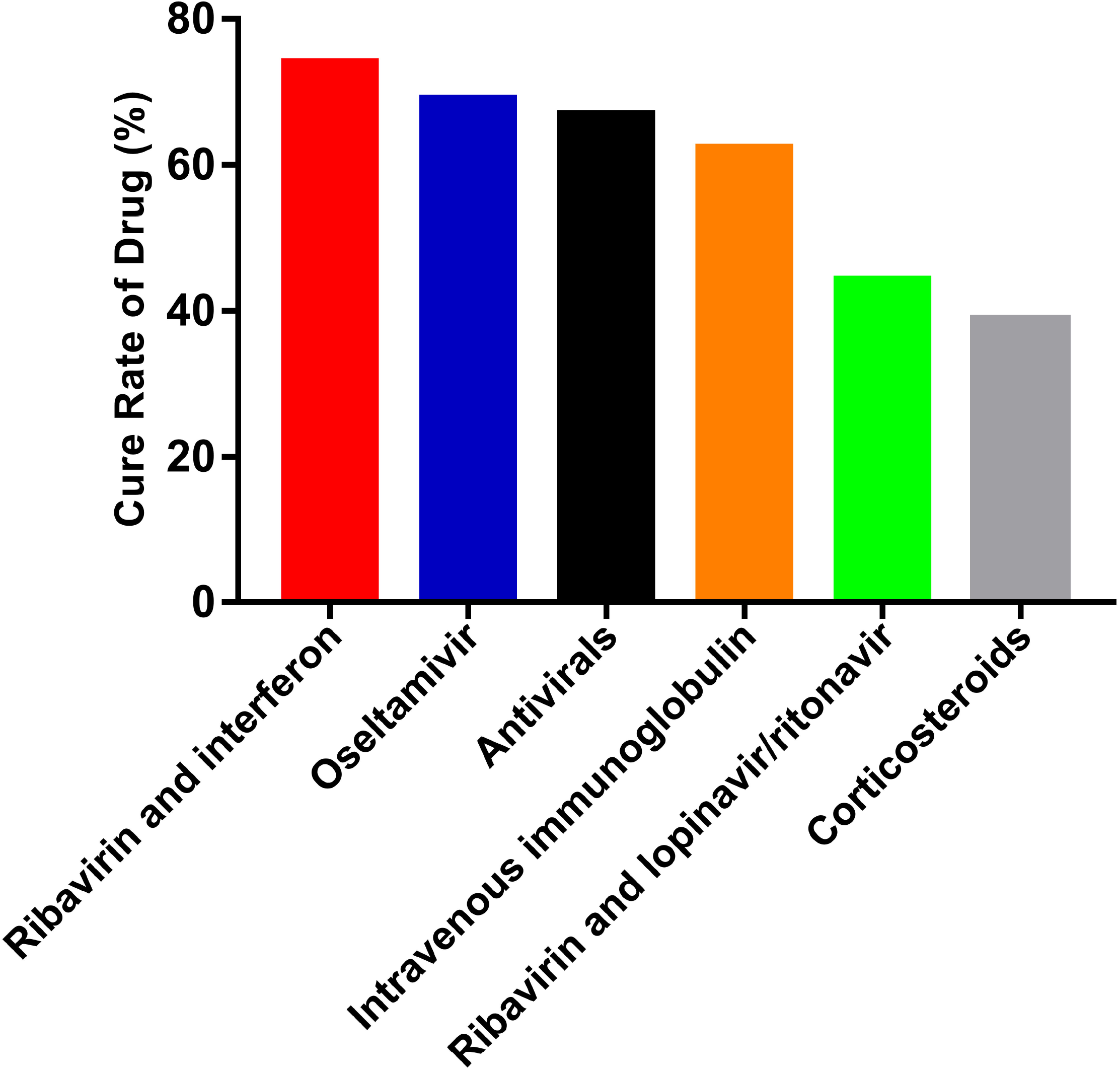

## Discussion

Coronavirus is an important pathogen causing respiratory and intestinal infection. Of seven identified coronaviruses, the two very pathogenic viruses, SARS-COV and MERS-COV, cause severe ARDS and even acute respiratory failure. With a mortality rate of more than 10% and more than 35% respectively [43-44]. The four other human coronaviruses (HCoV-OC43, HCoV-229E, HCoV-NL63, HCoV-HKU1) only cause mild respiratory or intestinal infection, despite they have certain pathogenicity for infants, young children and the elderly with weakened immune systems [45-46]. The newest one is COVID-19. The sequencing analysis has indicated that COVID-19, like SARS-COV and MERS-COV, belongs to β-coronavirus. Both SARS-COV and MERS-COV originated in bats, but the origin of COVID-19 remains to be further investigated. Although recent studies have reported the clinical and laboratory features of COVID-19, systematic comparsion between COVID-19 and MERS-COV has yet been performed. Therefore, the aim of this study is to perform the first systematic review to compare epidemiological, clinical and laboratory characteristics of COVID-19 and MERS-COV populations. Our results suggested that fever, cough and generalised weakness and myalgia were main clinical manifestations of both COVID-19 and MERS-COV, whereas ARDS was main complication. Compared with MERS-COV population, COVID-19 population had a less incubation time and lower rate of ICU admission, discharge and fatality.

Similarities of clinical characteristics between COVID-19 and MERS-COV have been found. In the current study, the majority of COVID-19 patients presented with fever, cough and generalised weakness and myalgia, which shows some resemblances to MERS-COV infection. Moreover, both COVID-19 and MERS-COV patients hardly developed upper respiratory tract infection such as rhinorrhoea or pharyngalgia, indicating that their target cells might be located in lower respiratory. However, 11.6% of patients with COVID-19 infection had diarrhoea or anorexia, and only 4.7% of patients with MERS-COV infection had this symptom. Thus, Faces and urine samples should be tested to exclude a potential alternative way of transmission that is unknown to the present.

In addition, we found that the number of males is more than that of females in either COVID-19 or MERS-COV population. The possible reason of reduced susceptibility of females to viral infection is that females have a lot of X chromosome and estrogen that are vital components in development of innate and adaptive immunity [47]. Meanwhile, numbers of patients with COVID-19 infection had chronic comorbidities, mainly hypertension, diabetes and cardiovascular disease, which is similar to MERS-COV population. Those results indicate that older adult males with chronic underlying disease might be more susceptibility to COVID-19 or MERS-COV.

In terms of laboratory testing, reduced lymphocytes and increased CRP were found in both COVID-19 and MERS-COV patients. This result indicates that COVID-19 might be associated with cellular immune response, mainly act on lymphocytes like MERS-COV does [48]. The cells infected by viruses induce the release of numbers of pro-inflammatory cytokines and inflammation storm in the body. Moreover, increased cytokines might make damage to related organs such as liver [49]. Our results demonstrated that abnormal value of AST was found in MERS-COV population, but not in COVID-19 population. The possible reason is that the follow-up time of COVID-19 population was too short, and the liver might remain to be in compensatory stage. For this result, a long follow-up time study is urgently needed. On the other hand, our results suggested that MERS-COV population had a higher rate of ICU admission and fatality than COVID-19 population, indicating that compared with MERS-COV, COVID-19 has less toxic and more easily cured. However, lower discharge rate was found in COVID population than in MERS-COV population. The possible explanation is that most of COVID-19 patients remained to be hospitalized at the time of manuscript submission, and data on those patients could not be obtained in time. Thus, careful interpretation is urged for this result.

Up to now, no effective strategy has been found for treatment of COVID-19 infection [50]. Currently, the measure to COVID-19 is to control the source of the infection; taking personal protective works to reduce the risk of transmission; and early diagnosing, isolating and supportive treating for confirmed patients. In the present study, our systematic results of cure rate of drug for MERS-COV infection indicated that ribavirin and interferon, oseltamivir, antivirals and intravenous immunoglobulin all had been effective for MERS-COV infection, with the cure rate of those drugs was 74.2%, 69.2%, 67.1% and 62.5% respectively. Thus, we assume that those drugs might also be effective at COVID-19 infection. However, further studies are needed to confirm this idea.

This study has several limitations. First, many patients infected by COVID-19 remained to be hospitalized at the time of manuscript submission, leading to the unavailable of some data. Second, the follow-up time of COVID-19 population is too short to get related data from long-term observations of this disease. Third, finding of statistical tests and p values between COVID-19 and MERS-COV populations should be interpreted with caution. Fourth, the number of MERS-COV patients treated by drugs is small, careful understanding is needed for the cure rate of drug for this disease. Finally, as COVID-19 is still developing around the world and remains to have many unknowns, the results of this study are staged and need to be carefully understood. More large-sample, multicentre, high-quality research should be performed to update this study.

## Conclusion

Our systematic review reveals that main clinical manifestations of both COVID-19 and MERS-COV populations are fever, cough and generalised weakness and myalgia. ARDS is main complication of both two populations. COVID-19 population has a shorter incubation time and lower rate of ICU admission, discharge and fatality compared with MRES-COV population.

## Data Availability

All data, graph, and model generated or used during the study appear in the submitted article

## Acknowledgments

This study was supported by grants from the National Natural Science Foundation of China (Nos.31970862) and Guangzhou Municipal Science and Technology project (Nos. 201803010001, Prof. Zhi-Zhong Li)

